# How should horizon scanning studies be reported? Developing a checklist of standard items

**DOI:** 10.1101/2025.10.23.25338620

**Authors:** Sonia Garcia Gonzalez-Moral, Meg Fairweather, Sarah Khan, Ross Fairbairn, Gill Norman

**Affiliations:** NIHR Innovation Observatory at Population Health Sciences Institute, Newcastle University

## Abstract

**Introduction:** Horizon-scanning identifies weak signals of innovation to anticipate future developments, providing strategic value for healthcare decision-making. Unlike evidence synthesis, it addresses emerging and uncertain areas but lacks standardised reporting, limiting transparency, consistency, and impact. Inconsistent terminology and poorly described methods hinder comparability and uptake. This study aimed to develop a prototype reporting checklist and glossary to support structured, transparent, and reproducible reporting of horizon-scanning methods in healthcare innovation.

**Methods:** A multidisciplinary working group of horizon-scanning, evidence synthesis, and information specialists was convened at the NIHR Innovation Observatory, the UK national horizon-scanning center, in May 2024. Using PRISMA-ScR as a framework, four workshops were held to adapt relevant reporting items and build a prototype checklist. Internal validation was conducted on 17 eligible Innovation Observatory reports (2017–2024), with scoring to assess item coverage. Items were then refined, classified as mandatory or optional, and finalised through consensus.

**Results:** Of 46 outputs screened, 17 met inclusion criteria. Fifteen checklist items achieved ≥50% coverage; 20 scored <50%. The final 35-item checklist (31 single and 4 multi-part items) includes 28 mandatory and 7 optional items. Four novel components were introduced: interest holder description, HIP-D/I scope framework, technology characteristics, and integration of the PESTLE framework.

**Discussion and recommendations:** This is the first checklist to standardise reporting in health technology horizon-scanning. While this checklist prototype has been developed internally, external validation involving multidisciplinary experts via Delphi and a scoping review are planned. Its adoption can enhance transparency, reproducibility, and strategic impact, strengthening this methodological field.

## Introduction

Horizon-scanning refers to the structured identification and analysis of early signals of innovation, enabling anticipation of future developments. It belongs to the broader field of Futures studies, which is concerned with exploring, modelling, and preparing for alternative future scenarios. Although horizon-scanning has been widely adopted across diverse sectors, its application in the medical sciences is relatively recent.(Meskó et al., 2024) The Innovation Observatory uses horizon-scanning to systematically detect signs of innovative healthcare technologies and support future decision-making processes about those.(Innovation Observatory, 2025)

Unlike evidence synthesis, understood as the process of systematically identifying, appraising, and integrating findings from multiple studies on a specific topic to generate comprehensive, reliable and policy or practice relevant conclusions,(Higgins JPT et al., 2024), Horizon-scanning focuses on the systematic process of detecting and collecting early signs and weak signals of potentially important developments which, in this context, they may be interpreted as innovative healthcare technologies.(Amanatidou et al., 2012) However, horizon-scanning currently lacks standardised reporting guidance, particularly for key aspects of the horizon-scanning process such as the identification, selection, and prioritisation of emerging health technologies. Furthermore, a recent review of horizon-scanning practices in medical technologies, diagnostics, and digital health recommended the use of dedicated reporting standards in light of the inconsistent use of terminology within the field.(Garcia Gonzalez-Moral et al., 2023) This lack of clarity impedes the dissemination and utility of horizon-scanning outputs—such as published reports and research articles—and contributes to research inefficiencies, including duplication and underutilisation of findings.(Glasziou & Chalmers, 2018)

Horizon-scanning is more likely to be used to widely explore the ‘unknowns’, being those more or less certain and definitively less-defined, on emerging areas of innovation across diverse domains. Using the Rumsfeld matrix as a conceptual framework and using it to compare with other well established methodologies such as evidence synthesis, we could argue that while evidence synthesis is akin to examining the visible tip of an iceberg (‘known knowns’), horizon-scanning focuses on what lies beneath the surface (‘unknown knowns’ and ‘known unknowns’).(Garvey et al., 2022)

While horizon-scanning is recognised as a methodological approach in its own right, it requires clear principles and structured processes to ensure rigour and consistency. Despite its growing use in health and policy contexts, the field remains underserved by methodological frameworks, reporting standards, or dedicated guidance. This absence of consensus risks limiting the transparency, comparability, and overall impact of horizon-scanning outputs. Reporting guidelines specify a minimum set of elements that should be included in research publications and have been shown to enhance methodological transparency and promote the use of research findings.(Tricco, 2018) To our knowledge, no standard reporting guidelines currently exist to support the transparent and consistent documentation of horizon-scanning activities in the healthcare domain. Given the increasing role of horizon-scanning in healthcare decision-making—both in the UK and internationally—this study aims to address this gap by developing the first reporting checklist for horizon-scanning methods in medical innovation.

### Aim and objectives

This study aims to initiate the standardisation of reporting practices for horizon-scanning methods and tools, thereby enhancing the transparency and reproducibility of such approaches.

Our objectives include the development of a conceptual framework for horizon-scanning research question formulation and query translation and the development of a preliminary checklist specifying essential and optional elements that should be transparently reported in horizon-scanning outputs, and a glossary to support consistent interpretation and reduce ambiguity in terminology. Further work will support finalisation of the checklist, incorporating the experience of those using this preliminary version.

### Scope of this checklist

This checklist is intended for application to horizon-scanning outputs including both articles published in academic journals and reports published on institutional websites. The checklist is developed to provide detailed and transparent information about the application of horizon-scanning methods to medical health technologies. Intended users of this checklist include authors of such reports, peer-reviewers of manuscripts submitted to journals for publication as well as journal editors when assessing such manuscripts and methods.

## Methods

A working group comprised of horizon-scanning, evidence synthesis, and information specialists was formed at the NIHR Innovation Observatory in May 2024. Participants for the working group were identified based on expertise in horizon-scanning, evidence synthesis and information science. Four workshops that spanned two hours each were scheduled between May and October 2024 to outline objectives, refine methods, and build the checklist.

In the pre-workshop meeting, the group set its goals and procedures. Given the similarity of broad scoping reviews with horizon-scanning methods, we decided to use the PRISMA extension for scoping reviews (PRISMA-ScR) as a framework to develop a checklist for reporting items relevant to horizon-scanning reports.(Tricco, 2018) During the first and second workshops, the group assessed the relevance of each PRISMA-ScR checklist item against the methodological and topic characteristics of horizon-scanning reports and produced a modified checklist prototype for horizon-scanning studies.

For the internal validation of the prototype, we applied the checklist to outputs produced by the Innovation Observatory between 2017 and 2024. Each team member in the working group was assigned a set of randomly selected reports to screen for suitability and, if applicable, score against our checklist prototype those reports that used horizon-scanning methods for the identification of innovative healthcare technologies. Following the screening process, 17 reports were deemed suitable for scoring. The rest were excluded as they were not horizon-scanning reports. Each checklist item was either marked as ‘yes’ (included), ‘no’ (not included), or not applicable (N/A) if the item was not relevant to this specific paper. Following completion of the scoring exercise, one team member (MF) combined the scores for each item and calculated a final percentage score by dividing the score by the maximum possible score (N/A items were not included in the maximum score).

**Figure 1.**
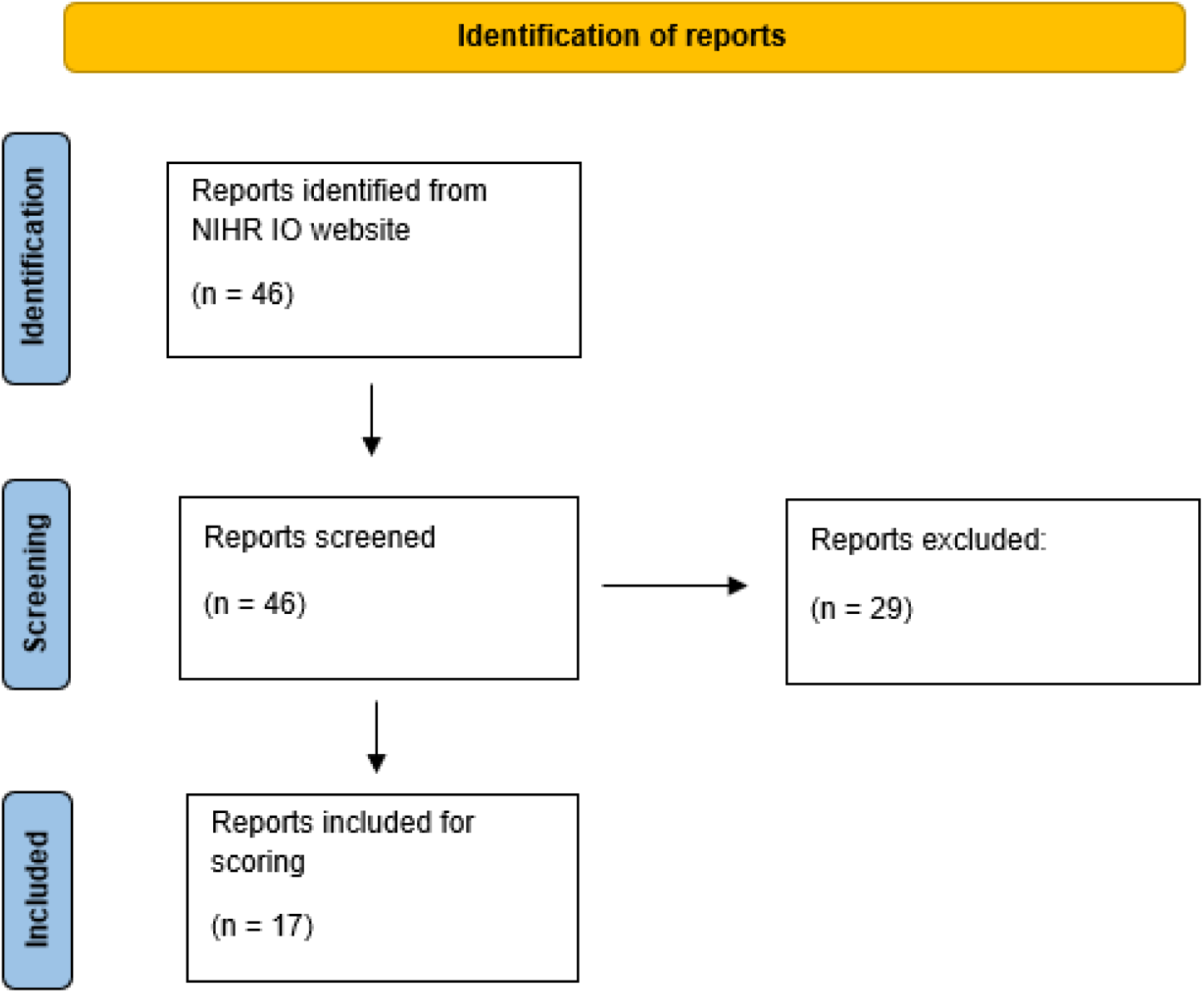
PRISMA flow diagram depicting included and excluded reports

In the third and fourth workshops, the working group reviewed the scores, proposed additional customisation of items to address issues relevant to horizon-scanning based on the scoring exercise and achieved consensus on whether the items should be made mandatory or optional. The resulting checking prototype was uploaded to Open Science Framework for dissemination.

## Results

A total of forty-six outputs published between 2017 and 2024 were downloaded from the NIHR Innovation Observatory website by a team member (OW). Seventeen horizon-scanning reports met the criteria for inclusion in this assessment and were assessed against the checklist. The remaining reports were not horizon-scanning reports and were not suitable for this assessment, the reasons for exclusion of these were: only an abstract, a methods paper or a presentation (not a report).

Fifteen items achieved a final score between 50 and 100% while 20 items were scored under 50%. Table 1 presents a breakdown of these items by the total final score.

**Table 1.**
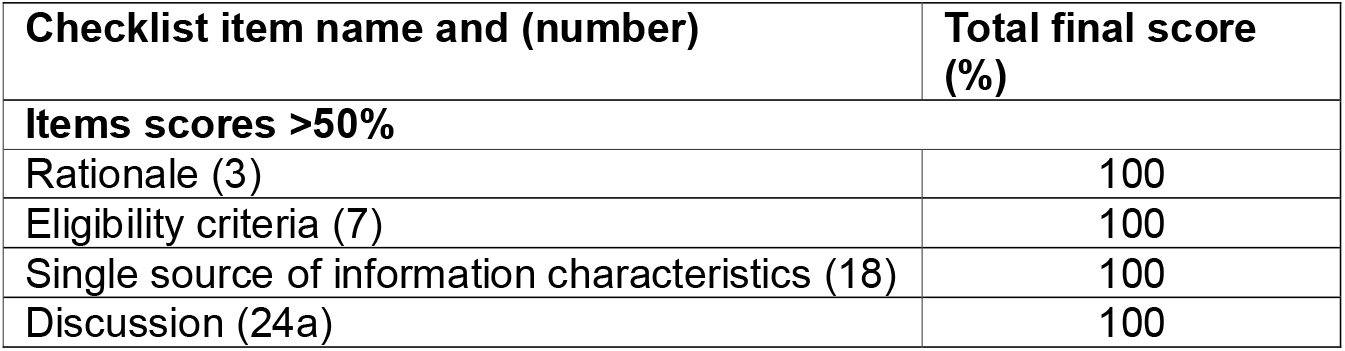
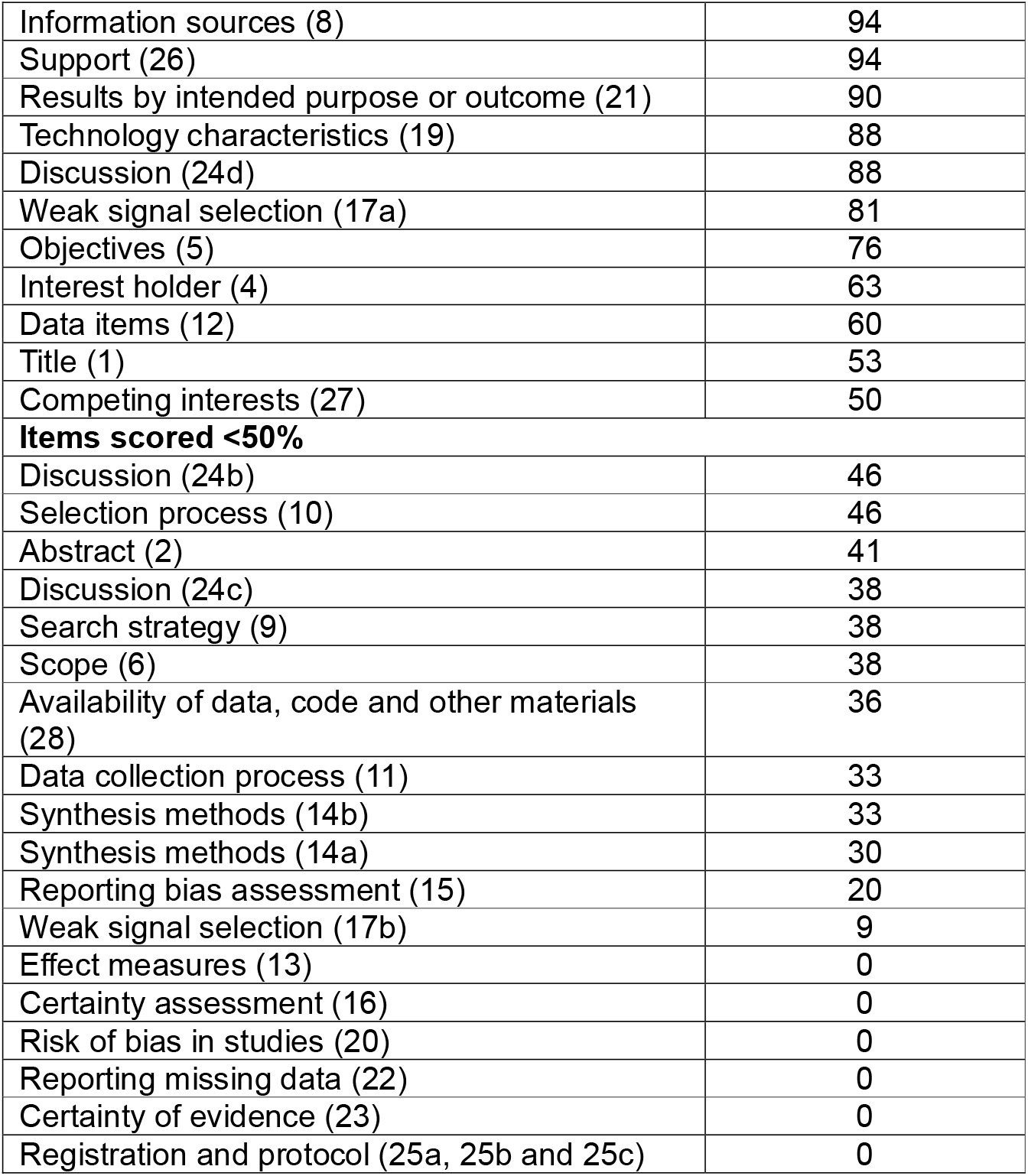
Scoring checklist items by number of times the item was present in the horizon-scanning reports assessed.

| Checklist item name and (number) | Total final score (%) |
| --- | --- |
| **Items scores >50%** |  |
| Rationale (3) | 100 |
| Eligibility criteria (7) | 100 |
| Single source of information characteristics (18) | 100 |
| Discussion (24a) | 100 |
| Information sources (8) | 94 |
| Support (26) | 94 |
| Results by intended purpose or outcome (21) | 90 |
| Technology characteristics (19) | 88 |
| Discussion (24d) | 88 |
| Weak signal selection (17a) | 81 |
| Objectives (5) | 76 |
| Interest holder (4) | 63 |
| Data items (12) | 60 |
| Title (1) | 53 |
| Competing interests (27) | 50 |
| **Items scored <50%** |  |
| Discussion (24b) | 46 |
| Selection process (10) | 46 |
| Abstract (2) | 41 |
| Discussion (24c) | 38 |
| Search strategy (9) | 38 |
| Scope (6) | 38 |
| Availability of data, code and other materials (28) | 36 |
| Data collection process (11) | 33 |
| Synthesis methods (14b) | 33 |
| Synthesis methods (14a) | 30 |
| Reporting bias assessment (15) | 20 |
| Weak signal selection (17b) | 9 |
| Effect measures (13) | 0 |
| Certainty assessment (16) | 0 |
| Risk of bias in studies (20) | 0 |
| Reporting missing data (22) | 0 |
| Certainty of evidence (23) | 0 |
| Registration and protocol (25a, 25b and 25c) | 0 |

Following, based on the working group members’ experience and in line with the objectives of this study, the items were rated by agreement as optional or mandatory. An optional item meant that the information required was not always possible to disclose due to a number of reasons that include technology stage of development, stakeholder agreements, availability of the information or relevancy of the item in the context of the horizon-scan scope. These items start with ‘If appropriate’ statement to indicate their optionality. A mandatory item was deemed of the upmost importance for the conduct of horizon-scanning methods and interpretation of results. These needed to be present at all times in order to achieve transparency in the retrieval and filtration of the signals, replicability of methods, avoid bias in the results, accountability and impact and overall increased rigour of the final horizon-scan output.

The resulting checklist contains 35 items (31 single items plus four that include multiple sub-items) of which 28 were deemed mandatory and seven optional (items 15, 16, 17b, 19, 20, 21 and 23). Some subsections such as “Synthesis methods” (items 14a and 14b) in the Methods section and “Weak signal selection” (items 17a and 17b) in the Results section present more than one item per section with only item 17b considered optional. The sections “Discussion” (items 24a to 24d) and “Registration and protocol” (items 25a to 25c) also include multiple sub-items and are all considered mandatory.

This checklist includes four new essential components not found in PRISMA-ScR, these are: “Interest holder description” item 4 in the Introduction, “Scope” item 6 in the Methods, “Technology characteristics” item 19 in the Results sections and the introduction of the PESTLE (Policy, Economy, Societal, Technological, Legal and Environmental) framework in item 24a in the Discussion section.

### Interest holder description

In the context of health technology horizon-scanning, an interest holder (often called a stakeholder) refers to any individual, group, or organization that has an interest in or may be affected by the development, adoption, use, or regulation of a health technology.

Key types of interest holders in this context may be:

1. Patients and Patient Advocacy Groups – interested in access, effectiveness, and safety of new and innovative health technologies.
2. Clinicians and Healthcare Providers – interested in clinical utility, integration into practice, and outcomes.
3. Health Technology Developers/Manufacturers – concerned with market potential, regulation, and innovation uptake.
4. Regulators and Health Authorities – interested in safety, efficacy, and compliance with regulatory standards or the early identification of gaps in current regulatory frameworks.
5. Policy Makers and Health System Planners – focused on broader system impact and public health implications.
6. Researchers and Academics – interested in evidence generation, gaps, and methodological development.
7. Health Technology Assessment (HTA) bodies – use horizon-scanning to inform evaluation and prioritisation.

Interest holders contribute by identifying unmet needs and emerging technologies; understanding key eligibility criteria based on needs or gaps; informing prioritisation criteria; providing early feedback on clinical relevance and implementation barriers; and supporting evidence appraisal and decision-making. Their involvement ensures that the scanning process is contextually relevant, anticipatory, and aligned with real-world health system priorities.

### Horizon scan scope

HIP-D/I Framework for scoping health technology horizon scans.

The HIP-D/I framework provides a structured approach to define the scope of horizon-scanning projects targeting innovative healthcare technologies. It consists of five key dimensions:

- **Horizon**: Categorises technologies based on their developmental proximity to market launch—emerging (early-stage concepts), transitional (in pre-commercial or pilot phases), and imminent (near-market entry and post-market surveillance). This helps determine the level of readiness and the timeframe for potential impact.
- **Innovation**: Focuses on identifying what is novel or disruptive about the technology compared to current standards of care. This includes technological, procedural, or conceptual innovations that may transform clinical practice or health system delivery.
- **Population**: Specifies the target patient group or population segment the technology is intended to benefit. Clarifying the population helps align horizon-scanning priorities with unmet clinical needs and equity considerations.
- **Data sources**: Emphasises the importance of identifying weak signals of innovation. Given the developmental stage of many technologies, relevant data may be found in non-traditional sources such as preprints, patents, clinical trial registries, venture capital investments, and early-stage conference presentations. Data sources and horizon are intrinsically related.
- **Interest holder:** As defined above, they provide a contextual element to the results of the horizon-scanning, enabling greater and immediate impact.

Together, these five dimensions support a systematic scoping process that enables transparent and reproducible methods for the identification and prioritisation of transformative health technologies.

### Technology characteristics

In horizon-scanning studies of healthcare innovation, providing a detailed description of the identified, filtered, and prioritised technologies is essential to ensure that outputs are meaningful and actionable for decision-makers. Unlike systematic reviews—where the unit of analysis is typically the study and its outcomes—horizon-scanning primarily focuses on the identification of innovative technologies and often uses the technology and its characteristics e.g. trial phase or technology readiness levels, as the primary unit of interest. Detailed technology descriptions capture critical attributes such as intended use, mechanism of action, stage of development, anticipated benefits, implementation challenges, and potential impact on health systems.

This level of detail serves several purposes: it supports informed prioritisation, facilitates future assessments (e.g., health technology assessment or regulatory review), enhances transparency and reproducibility, and enables strategic planning for health system readiness.

To align with these goals, the reporting checklist was adapted to shift the focus from evidence appraisal, as seen in PRISMA-ScR, to the detection of weak signals, in keeping with the purpose and definition of horizon-scanning.(Hines et al., 2019) Table 2 presents the prototype checklist, including item descriptions and corresponding document sections.

**Table 2.**
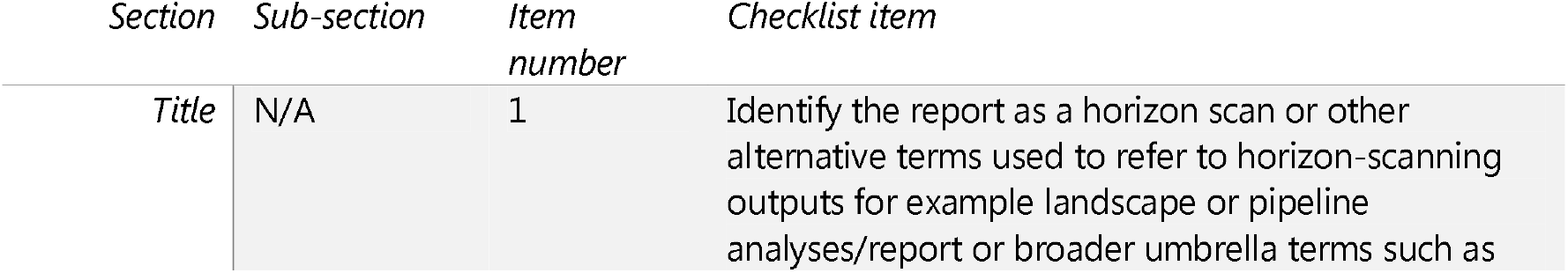
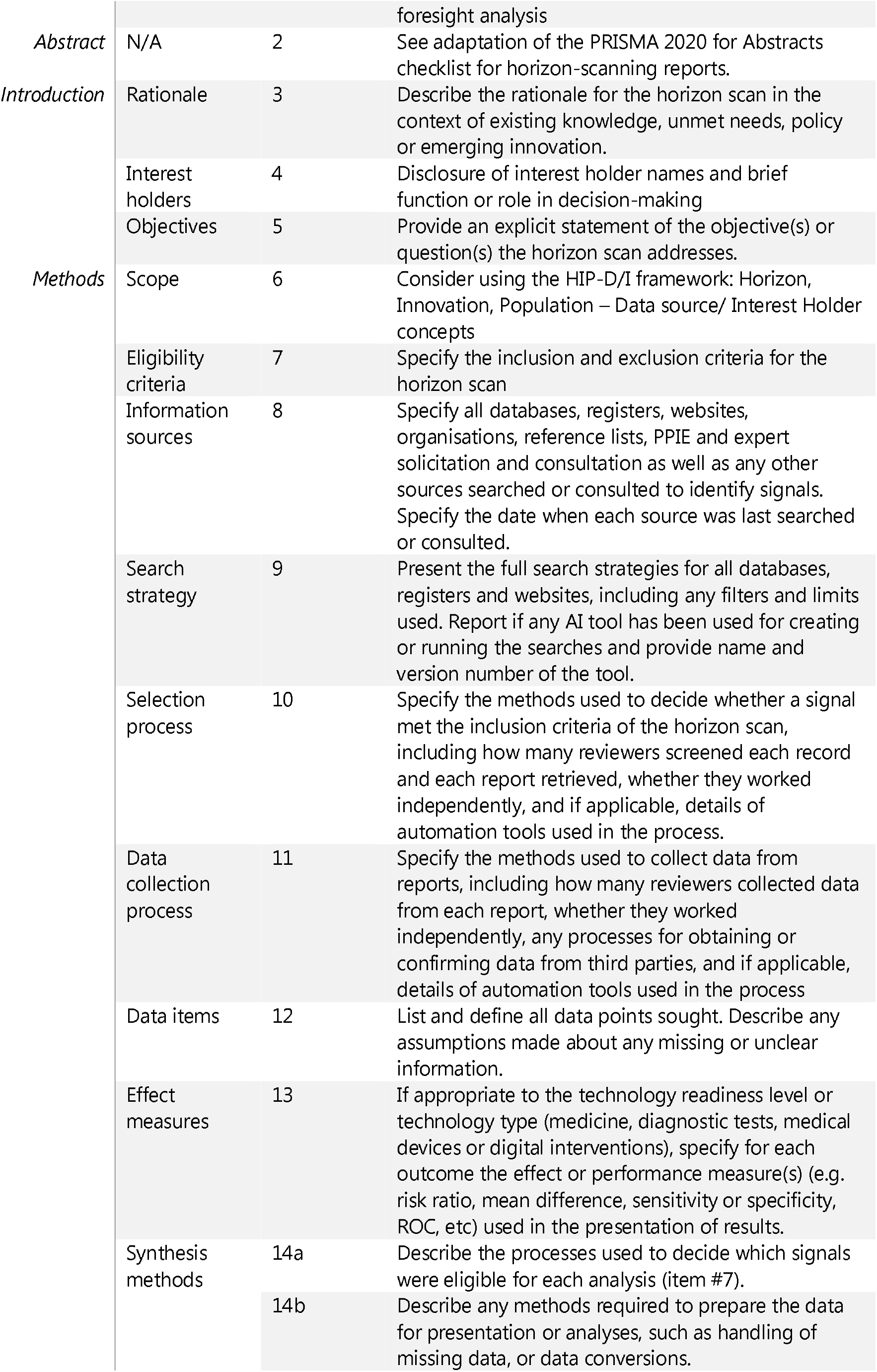
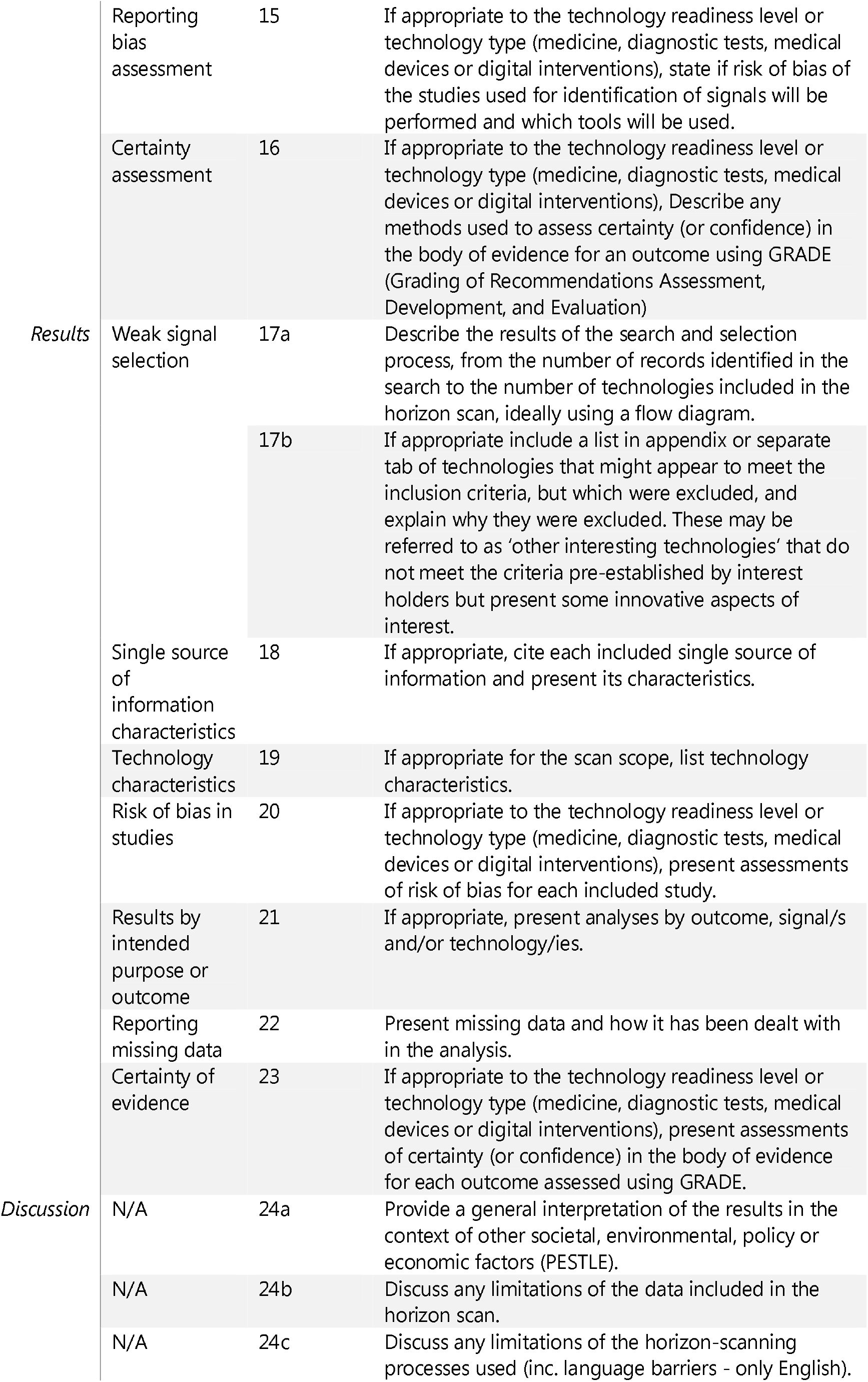
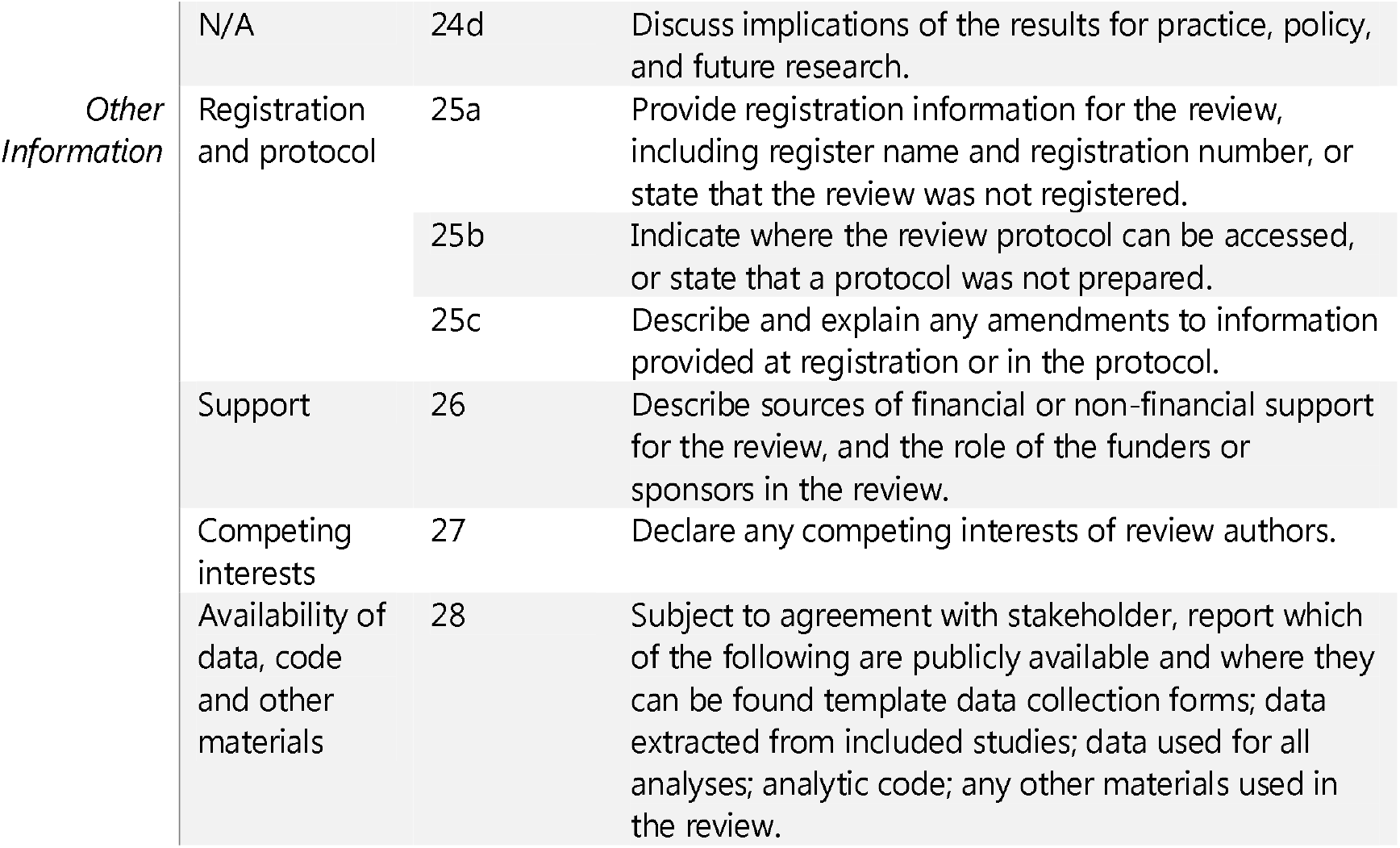
Final checklist items.

### PESTLE framework

Incorporating the PESTLE framework into the discussion section of a horizon-scan on emerging healthcare technologies enriches the report by situating innovations within a structured macro-environmental context, thereby highlighting how regulatory dynamics, economic constraints, social values, and environmental considerations intersect with technological development and diffusion. For instance, a strategic environmental scan of Iranian public hospitals demonstrated that PESTLE-guided analysis enabled the identification of external pressures—including economic sanctions, shifts in disease patterns, and governance issues—which informed adaptive strategies for equitable resource allocation and service delivery.(Pourmohammadi et al., 2020) Similarly, scoping reviews applying PESTLE-type analysis to electronic health record implementation revealed how political, economic, sociocultural, and legal dimensions shape adoption trajectories and reveal implementation challenges in low- and middle-income settings.(Mwogosi & Mambile, 2025) By framing horizon-scan findings within these six domains, authors can present a reproducible and comparative analytic lens that directly links emerging technologies to broader system drivers, aids in anticipating barriers and enablers to adoption, and enhances the strategic relevance of the report for policymakers, funders, and health system planners.

The full proposed checklist is shown in Table 2 and Appendix B.

The checklist is available to download from *Gonzalez-Moral SG, Khan SK, Fairbairn R, Williams O, Fairweather M, Norman G. Developing a checklist for transparent reporting of items in horizon-scanning studies [Internet]. OSF; 2025. Available from: osf*.*io/m39vw*. A full item by item description with text exerts as examples is provided in Appendix A. Appendix B provides the checklist form for completion at reporting writing stage. A glossary of terms for ease of interpretation of the checklist items is provided in Appendix C.

## Discussion

This study introduces what is, to the best of our knowledge, the first horizon-scanning checklist designed to enhance transparency, reproducibility, and methodological rigour in the reporting of health technology horizon-scanning studies. It presents the results of preliminary work, involving analysis of our own reports and in-depth discussions among expert practitioners within the horizon-scanning center (HSC). The checklist outlines identified key features that should be systematically reported to enable comprehensive interrogation and interpretation of intelligence derived from horizon-scanning. When assessed against the checklist, just over one-third of the items (15 of 35) achieved a final score above 50%, whereas the remaining 20 items were insufficiently reported. This variation in reporting highlights a lack of standardisation in current horizon-scanning outputs and demonstrates the need for a structured reporting framework. Six checklist items (13, 16, 20, 22, 23 and 25) received a score of 0, meaning they were not reported in any of the included reports assessed. Some of these items might not be relevant to all technology readiness levels and should not always be reported. For example, a horizon scan in the early horizon that uses patents and pre-clinical studies to identify trends and signals of innovative technologies will not present sufficient data on outcomes or the strength of the evidence to warrant risk of bias or certainty assessments. Furthermore, although it is strongly recommended that a horizon scan follows a robust, pre-agreed protocol, the highly strategic nature of some horizon-scanning makes a publicly available protocol unfeasible due to confidentiality agreements with stakeholders. Notably, many of these features are already represented in reports from the national HSC, highlighting both their relevance and feasibility. This finding demonstrates the need for more consistent and comprehensive reporting across all horizon-scanning studies to support full transparency, comparability, and utility of findings.

Key strengths observed across well-reported items included the consistent articulation of rationale, eligibility criteria, and technology characteristics—crucial elements for supporting decision-making. However, core methodological domains such as search strategy, selection process, synthesis methods, and discussion of limitations were frequently underreported or absent. These gaps limit the transparency and replicability of horizon-scanning studies, potentially reducing their utility for decision-makers involved in regulatory, reimbursement, or research prioritisation pathways.

To address these issues, the study team developed a prototype 35-item checklist (including multi-component sub-items), of which 28 were designated as mandatory and seven as optional. These designations were based on the expert consensus of internal team members and reflected both practical constraints and methodological requirements. It is worth stressing that the checklist introduces four essential components not found in existing reporting frameworks such as PRISMA-ScR: interest holder description, scope (HIP-D/I framework), technology characteristics and PESTLE framework. These additions are fundamental to horizon-scanning, which—unlike systematic reviews—uses the technology as the unit of analysis rather than the study or its outcomes. A detailed description of the identified, filtered, and prioritised technologies is critical to enable strategic planning, future assessments, and timely decision-making.

Furthermore, this checklist reflects the exploratory and anticipatory nature of horizon-scanning. The emphasis is on the detection of weak signals, in alignment with the goals of identifying transformative technologies ahead of market entry. This reframing supports the unique objectives of horizon-scanning, particularly where conventional data may be unavailable and insights must be drawn from diverse, and sometimes non-peer-reviewed sources such as clinical trial registries, patent databases, preprints, and expert consultation.

Horizon-scanning is increasingly used by a range of stakeholders—regulators, HTA bodies, policy-makers, clinicians, and technology developers—as a forward-looking, market-intelligence and decision-support tool.(McCool K, 2025; 2025; 2025; *Seeing the Big Picture: The Role of Horizon Scanning and Strategic Foresight in SMEs*, 2025) Given the variability in how horizon-scanning is defined and implemented across organisations, the proposed checklist may provide a common foundation for improving the transparency, comparability, and rigour of horizon-scanning practices in healthcare innovation.

Our study has several limitations. First, the development of the checklist prototype relied exclusively on internal experts from the NIHR Innovation Observatory. While the team possesses extensive experience in conducting horizon scans across a variety of health technologies, stakeholders, and time horizons, we recognise that this internal scope may limit the generalisability of findings. To address this, we plan to undertake a formal external validation using a scoping review and a multi-stage adapted Delphi process to achieve broader consensus on item relevance, structure, and applicability across diverse horizon-scanning contexts. Second, the checklist scoring was based on a relatively small sample of 17 reports published since 2017. Given the niche nature of horizon-scanning in healthcare, this limited sample size may have influenced the ranking of optional versus mandatory items. Future application of the checklist to a larger set of outputs will help mitigate this constraint and may refine item prioritisation.

### Conclusion and next steps

This study presents the development of a prototype checklist designed to improve the transparency, consistency, and methodological clarity of horizon-scanning reports for healthcare innovation. The checklist was created in response to a recognised gap in reporting standards,(Garcia Gonzalez-Moral et al., 2023) drawing on elements from the PRISMA-ScR framework(Tricco, 2018) to accommodate the broad and exploratory nature of horizon-scanning studies similar to those of broad scoping reviews.(Akartuna et al., 2024)

The checklist provides a structured foundation for reporting horizon-scanning outputs, facilitating more robust and reproducible methods, and supporting stakeholder engagement in early decision-making processes. Its adoption has the potential to contribute to the standardisation of horizon-scanning practices across organisations, enhance the strategic value of these outputs in innovation management and provide horizon-scanning methods of greater scientific rigour.

Next steps include conducting an external validation using a multi-stage Delphi process, following a rapid scoping review, involving a diverse group of horizon-scanning experts and end-users. This will help refine and potentially widen the checklist items, confirm their relevance across different settings, and promote broader adoption. If widely implemented, this checklist can serve as a key tool for strengthening the scientific basis of horizon-scanning and advancing its role in health policy and innovation planning.

## Supporting information

Checklist explanation with examples

Horizon Scanning checklist

## Data Availability

All data produced are available online in the Open Science Framework repository.

https://osf.io/m39vw/overview

## Acknowledgements

The authors would like to acknowledge Oleta Williams for her valuable contribution as an information specialist, particularly for her input into search strategy development and methodological discussions that informed this research.

This study/project is funded by the National Institute for Health and Care Research (NIHR) [NIHR IO/project reference HSRIC-2016-10009]. The views expressed are those of the author(s) and not necessarily those of the NIHR or the Department of Health and Social Care.

## References

Akartuna, E. A., Johnson, S. D., & Thornton, A. (2024). Enhancing the horizon scanning utility of futures-oriented systematic and scoping reviews. Futures, 158, :103340. 10.1016/j.futures.2024.103340

Amanatidou, E., Butter, M., Carabias, V., Könnölä, T., Leis, M., Saritas, O., Schaper-Rinkel, P., & Van Rij, V. (2012). On concepts and methods in horizon scanning: Lessons from initiating policy dialogues on emerging issues. Science and Public Policy, 39(2), :208–221.

Garcia Gonzalez-Moral, S., Beyer, F. R., Oyewole, A. O., Richmond, C., Wainwright, L., & Craig, D. (2023). Looking at the fringes of MedTech innovation: a mapping review of horizon scanning and foresight methods. BMJ Open, 13(9), :e073730. 10.1136/bmjopen-2023-073730

Garvey, B., Humzah, D., & Le Roux, S. (2022). Uncertainty deconstructed: A guidebook for decission support practitioners. (Springer, Ed.). 10.1007/978-3-031-08007-4

Glasziou, P., & Chalmers, I. (2018). Research waste is still a scandal—an essay by Paul Glasziou and Iain Chalmers. BMJ, 363, k4645. 10.1136/bmj.k4645

Higgins JPT, Thomas J, Chandler J, Cumpston M, Li T, Page MJ, & (editors)., W. V. (2024). Cochrane Handbook for Systematic Reviews of Interventions version 6.5 (updated August 2024). https://www.training.cochrane.org/handbook.

Hines, P., Hiu Yu, L., Guy, R. H., Brand, A., & Papaluca-Amati, M. (2019). Scanning the horizon: a systematic literature review of methodologies. BMJ Open, 9(5), :e026764. 10.1136/bmjopen-2018-026764

Innovation Observatory. (2025). A world leading horizon scanning facility. https://io.nihr.ac.uk/

McCool K. (2025). UK to accelerate patient access to medicines under new MHRA–NICE joint approach. https://becarispublishing.com/digital-content/blog-post/uk-accelerate-patient-access-medicines-under-new-mhra-nice-joint-approach

Meskó, B., Kristóf, T., Dhunnoo, P., Árvai, N., & Katonai, G. (2024). Exploring the Need for Medical Futures Studies: Insights From a Scoping Review of Health Care Foresight [Review]. J Med Internet Res, 26, :e57148. 10.2196/57148

Mwogosi, A., & Mambile, C. (2025). Applying PEST analysis to evaluate the adoption of electronic health records in Tanzanian healthcare: A scoping review. Digit Health, 11, :20552076251334029. 10.1177/20552076251334029

NHS. (2025). SPS Horizon Scanning. https://www.sps.nhs.uk/articles/sps-horizon-scanning/

NHS England. (2025). Horizon scanning and demand signalling. https://www.england.nhs.uk/aac/what-we-do/demand-signalling/

Pourmohammadi, K., Bastani, P., Shojaei, P., Hatam, N., & Salehi, A. (2020). A comprehensive environmental scanning and strategic analysis of Iranian Public Hospitals: a prospective approach. BMC Research Notes, 13(1), :179. 10.1186/s13104-020-05002-8

Seeing the Big Picture: The Role of Horizon Scanning and Strategic Foresight in SMEs. (2025). https://www.future-finance.tech/post/seeing-the-big-picture-the-role-of-horizon-scanning-and-strategic-foresight-in-smes

Tricco, A., Lillie, E, Zarin, W, O’Brien, KK, Colquhoun, H, Levac, D, Moher, D, Peters, MD, Horsley, T, Weeks, L, Hempel, S et al. (2018). PRISMA Extension for Scoping Reviews (PRISMA-ScR): Checklist and Explanation. Annals of Internal Medicine, 169(7), :467–473. 10.7326/m18-0850%m30178033

