## Supplementary material for "How should horizon scanning studies be reported? Developing a checklist of standard items": Checklist explanation with examples

### Appendix A. Itemised description of the checklist

**Title**

**Number of items in this section: 1**

**Item 1 | Title: *Identify report as a horizon-scanning or equivalent term in title* | Mandatory | Modified from PRISMA-ScR**

*Explanation:* Depending on the context and objectives, alternative terms can also be used to maximise specificity, such as ‘landscape analysis’, ‘pipeline analysis/report’, ‘trend analysis’, ‘weak signal detection’, or ‘foresight analysis’.

*Rationale:* Clearly identifying the report as a horizon scan ensures that it reaches the appropriate audience and maintains transparency from the outset.

*Example:* *“Interim Horizon-scanning Report: Identification of Paediatric Neurological Trauma Technologies”(Oparah C et al., 2023)*

**Abstract**

**Number of items in this section: 1**

**Item 2 | Abstract*: See adaptation of the PRISMA 2020 for Abstracts checklist* | Mandatory | Modified from PRISMA-ScR**

*Explanation:* Some adaptations have been made to the PRISMA 2020 for abstracts checklist. For example, while risk of bias is more relevant for late-stage technologies, it may not be relevant for all horizon scans and can be considered optional (this is expanded on in the results section below). For the results part of the abstract, focus on summarising key signals, technologies, or trends, rather than traditional study outcomes.

*Example: (results section in abstract) “1163 records were identified through searching ScanMedicine, and 136 trials were included in the final data extraction and mapping process. The most frequently investigated faecal incontinence intervention topics were complementary therapies (n=17, 12.4%); electrical stimulation (n=13, 9.5%); pelvic floor muscle training/biofeedback/sphincter exercises (n=13, 9.5%) and implanted sacral nerve stimulation (n=12, 8.7%). There was little evidence of new pharmaceutical technologies in development. Existing drugs are, however, being repurposed and trialled for the treatment of faecal incontinence (eg, linaclotide, colesevelam). Such repurposed drugs often have lower development costs, shorter timelines and report lower failure rates compared with new pharmaceutical products.”(Thomson et al., 2022)*

**Introduction**

**Number of items in this section: 3**

**Item 3 | Rationale*: Describe the rationale for the horizon scan in the context of existing knowledge, unmet needs, policy or emerging innovation |* Mandatory | Modified from PRISMA-ScR**

*Explanation:* Situate the report within the broader landscape or existing knowledge, unmet needs, policy, or emerging areas of innovation. Outline what is already known in the field, describe the gaps in current research and practice, and explain why these gaps are worth exploring. If possible, link the scan to current policy or policy priorities.

*Rationale:* Providing clear context at the beginning of the report helps the audience understand the purpose of the scan. This also ensures that the horizon-scan output is targeting a policy which support the study of its impact.

*Example: “The complexity of the pathology of COVID-19 has challenged the development of appropriate therapeutics … While certain therapies have shown benefit in a subset of the treatment population … the complexity of the disease necessitates the need to look beyond monotherapies and into combining independent treatments to increase therapeutic efficacy in a much wider population and across the disease pathway. There are several compelling reasons for exploring combination therapies for COVID-19 treatment: (i) different mechanisms … The use of multiple combination therapies, however, are known to be associated with increased risks in terms of safety, potential interactions and adverse effects, which may (at least partially) explain the low numbers currently being tested in trials for COVID-19.”(Akinbolade et al., 2022)*

**Item 4 | Interest holder: *Disclosure of interest holder name and brief function or role in decision-making* | Mandatory | New addition to checklist**

*Explanation:* This is strongly encouraged in agreement with the commissioner of the project. An interest holder is anyone who has an interest in a project, outcome, or decision. This can include stakeholders, who also have a direct influence or involvement in the decision-making process. Include the individuals full name or the name of the organisation they represent, as well as their current position or role within their organisation, what type of interest holder they are, and a brief description of what they contributed to the process. Also include any declared interests or potential conflicts if relevant. This could be broadened to cover other organisations that might be interested but have not requested the scan.

*Rationale:* Disclosing the name and role of each interest holder supports transparency and accountability. It clarifies who has had input into the process and helps readers interpret findings within the context of potential interests, expertise or influence. It also enhances the credibility of the scan by demonstrating balanced representation and helping to identify any conflicts of interest or biases that may need to be managed.

*Example: “As such, NHSx (via the Accelerated Access Collaborative) requested a horizon scan to ascertain the … Whilst interested in the application of AI broadly to health and social care (whole market), NHSx also specified 4 areas of particular interest … The technologies and intelligence identified in the horizon scan will be used further by NHSx to consider …”(Oyewole A et al., 2021)*

**Item 5 | Objectives: *Provide an explicit statement of the objective(s) or question(s) the horizon scan addresses* | Mandatory | | Modified from PRISMA-ScR**

*Explanation:* Clearly state the objectives or questions the horizon scan aims to address. It may be useful to link this back to the rationale.

*Rationale:* This ensures that readers clearly understand the scope and purpose of the scan, preventing misunderstanding or duplication and supporting accountability to interest holders and the public. It also strengthens the methodological integrity of the report by ensuring that the scan remains focused, structured, and transparent, while helping to minimise bias.

*Example: “Objectives 1) To complete a horizon scan of AI technologies across health and social care, including a targeted scan of mature AI technologies for scanning/medical imaging, oncology, cardiology, dermatology, and ophthalmology. 2) Summarise business intelligence on AI, with the focus on finding key themes and trends, including the activities of established firms such as Google.”(Oyewole A et al., 2021)*

**Methods**

Number of items in this section: 12

**Item 6: Describe the scope of the Horizon Scan** **| Mandatory | Modified from PICO**

*Explanation:* The HIP-D/I framework follows a modified PICO/S (Population, Intervention, Comparator, Outcome, Study type) framework which outlines four different potential components of an evidence-based health research questions. The HIP-D/I framework includes concepts such as Horizon; Innovation; Population and Data sources or Interest Holders. This framework represents a shift in focus from a systematic review perspective that follows a traditional PICO/S or some variation of the frameworks and aligns more with the purpose and the methods of horizon-scanning.

*Rationale:* Transparent reporting of the concepts that shape the scope of a horizon-scan project is vital for methodological rigour, reproducibility, and credibility. By clearly disclosing the frameworks, inclusion criteria, timeframes, and stakeholder inputs that informed scope decisions, authors enable critical appraisal, reduce interpretive bias, and foster comparability across studies.

*Example: “The scope of this horizon scan was to identify signals from clinical trials and funding sources relating to emerging GenAI-based technologies being developed for use in a clinical setting to improve patient care. GenAI-based technologies being developed to support healthcare systems or healthcare professionals rather than directly for patient care were not included in the scope of this scan. Technologies across all TRLs from initial technology concept through to technologies ready to be implemented in a clinical setting were included in the scope of this scan.”(Lanyi K et al., 2025)*

***Item 7: Specify the inclusion and exclusion criteria for the horizon scan* | Mandatory | Modified from PRISMA-ScR**

Explanation: Reporters are to ensure they clearly outline the inclusion and exclusion criteria for the horizon scan and must provide a rationale for why such criteria were selected. Specific restrictions such as date limits, language, location etc. should also be included as eligibility criteria along with a rationale for each restriction.

*Rationale:* Specifying inclusion and exclusion criteria in a horizon scan is mandatory because it establishes the boundaries of the scan in a transparent and reproducible way. Clear criteria ensure that the selection of technologies is systematic rather than arbitrary, which reduces bias, strengthens the validity of the findings, and allows stakeholders to understand why certain innovations were prioritised or omitted. This transparency not only enhances the credibility of the scan but also facilitates comparability across studies and supports informed decision-making by policymakers and planners.(Hines et al., 2019)

*Example (brief version): “Inclusion: Technologies must be regulated medical devices, digital health tools, or diagnostics under EU MDR (2017/755) or IVDR (2017/746). Must qualify as AI technologies, evidenced by: AI-specific keywords (e.g. “deep learning”), involvement of AI specialists, confirmation from secondary sources, description of machine learning methodology, data collection for AI training, or evaluation/development of AI-based devices (e.g. smartwatches, diagnostic tools, risk assessment/mitigation, or patient-level applications). Generative AI (GenAI) is included if explicitly referenced (e.g. GPT, transformers, neural networks) and demonstrated to generate new content for healthcare purposes, such as text automation, clinical decision support, diagnostic image enhancement, drug discovery, or personalised rehabilitation. Where unclear, secondary searches confirm GenAI use.*

*Exclusion: GenAI used only for training or education of healthcare professionals. GenAI applied at hospital/disaster management (system-level) rather than individual-patient level. Primary clinical research not involving patient-level outcomes. Patient information or education tools without direct impact on health outcomes. “Intelligent” technologies that collect enriched data but lack autonomous analysis. Predictive AI that only classifies or forecasts from inputs without generating new content.”(Lanyi K et al., 2025)*

***Item 8: Information Sources* | Mandatory | Modified from PRISMA-ScR**

Explanation: It is recommended that a comprehensive literature search is conducted which may include literature from bibliographic databases, clinical trials databases, news sites etc. All sources of information must be specified in this section, in addition to dates each source was searched and/or consulted to demonstrate relevancy of information used within the report. It is also recommended to describe the rationale for unconventional sources, methods or supplementary searching if necessary (for example, hand-searching key journals).

*Rationale:* Transparent disclosure of information sources in a horizon scan report is essential to ensure reproducibility, credibility, and methodological rigour. Clearly reporting databases, trial registries, websites, and other sources allows readers to understand how technologies were identified, reduces selection bias, and enables verification or replication of the search process. This practice aligns with PRISMA‑ScR guidelines, which emphasise comprehensive and transparent reporting of sources to enhance the trustworthiness and utility of scoping and horizon scan reviews.(Tricco, 2018)

*Example*: *“The Embase (Ovid) search strategy was adapted to other database interfaces and run in the Cochrane Database of Systematic Reviews, TRIP database, MEDLINE (Ovid) and Scopus. A supplementary search in PubMed for publications entered in the database since 2022 was run to cover the potential gaps generated by unindexed publications.”(Garcia Gonzalez-Moral et al., 2025)*

***Item 9: Search Strategy*** **| Mandatory | Unmodified from PRISMA-ScR**

*Explanation*: As per the PRISMA-ScR guidelines, it is recommended to provide full search strategies for all databases, registries and websites in addition to any limits or filters that were applied to ensure that the search is transparent and reproducible. Any AI or machine learning tools that were used to construct or run the search itself must also be reported with full details of software/tool name and version if applicable. Full strategies should be listed in the text, table or appendix. Details of who constructed and run the search (i.e. and Information Specialist) in addition to whether the strategy was peer reviewed should be documented.

*Rationale:* By fully reporting databases, search terms, limits, and inclusion criteria, others can assess the completeness and potential biases of the evidence base. This is critical since horizon scanning aims to identify emerging trends or innovations where omission or selective reporting could distort priority-setting and policy decisions. Clear documentation also facilitates updating and validation of the scan by other researchers or stakeholders.

*Example: “…a search strategy was designed in Embase (Ovid) by one information specialist (OW) and peer-reviewed by another information specialist (SGG-M). The search strategies consisted of a combination of keyword and free-text terms in title and abstract and keyword fields. We searched for (COPD OR Asthma) AND (devices OR diagnostics OR POC). No language or publication type limits were imposed. […] We used PubVenn, a search application that enables visualisation of PubMed searches through Venn diagrams and assists with rapid identification of relevant sets of papers. In addition, we used citation tracking options such as ‘cited by’ and ‘similar papers’ in PubMed to identify potentially relevant papers. We searched reference lists of known reviews to identify additional papers that included the technologies of interest. Additionally, we searched the Google and Apple app stores for digital apps for self-monitoring of asthma and COPD. We launched Google searches for known technologies and sifted results on screen. We also searched company and manufacturers’ websites for additional products in scope, checked the British Lung Foundation website for additional technologies and consulted with relevant stakeholders. All searches were undertaken iteratively and completed between December 2022 and March 2023. Preferred Reporting Items for Systematic reviews and Meta-Analyses Statement (PRISMA-S) extension has been followed for reporting the searching element of this review. Full database strategies are also included in online supplemental appendix 1, section 3.”(Garcia Gonzalez-Moral et al., 2025)*

***Item 10: Selection Process* | Mandatory | Modified from PRISMA-ScR**

*Explanation*: The full screening process must be described including number of screeners and whether any automated (AI or machine learning) tools or methods were used in the process, reporting software/tool name and version if applicable.

*Rationale:* Transparently reporting the selection process in a horizon scan of innovative technologies allows stakeholders to understand how and why certain technologies were prioritized over others. This reduces the risk of bias or perceived subjectivity, strengthens the credibility and legitimacy of the findings, and enables others to replicate, critique, or build upon the work. Clear documentation also supports accountability, particularly when horizon scan outputs are used to inform policy, investment, or research priorities.

Example: *“…two reviewers sifted and selected all relevant technologies from the identified included reviews and relevant evaluations. Sifting was undertaken by one reviewer, and a second reviewer checked queries and assessed the relevance of the selected technologies in line with the inclusion and exclusion criteria (online supplemental appendix 1, table 1). Any disagreements were resolved by discussion or involving a senior member of the team.”(Garcia Gonzalez-Moral et al., 2025)*

*“Each record was screened and tagged as either ‘include’ or ‘exclude’. Ten percent of all records were double screened to ensure consistency amongst the reviewers, the remaining records were distributed across the review team and single-screened. During single screening, any record where the screener had a query was marked as such and discussed amongst the full screening team to reach a consensus.”(Lanyi K et al., 2025)*

***Item 11: Data Collection Process* | Mandatory | Unmodified from PRISMA-ScR**

*Explanation*: The data collection process, also referred to as data extraction, involves the systematic capture of predefined data points from the included records. These data elements, established during protocol development, are selected to ensure consistency and relevance. Their purpose is to provide the necessary evidence base to support the robust assessment, selection, and prioritisation of technologies.

*Rationale:* Reporting the methods for data extraction in a horizon scan or scoping review is important because it shows how information about the identified technologies was systematically captured, organized, and interpreted. Transparent reporting allows others to judge the consistency, accuracy, and completeness of the extracted data, reduces the risk of selective reporting or bias, and supports reproducibility. It also ensures that the evidence base used to describe and compare emerging technologies is reliable and comparable, which is crucial for informing research, policy, or investment decisions.

*Example*: *“We designed a data extraction form that presented all prespecified data points needed to support decision-making and agreed with stakeholders at protocol stage. Some of these datapoints required further targeted searches on clinical trial registries such as ClinicalTrials.gov, additional bibliographic databases and company websites. These searches were mostly performed on company or product name and were undertaken by horizon scanning analysts. All additional information captured by these searches was added to the data extraction form.”(Garcia Gonzalez-Moral et al., 2025)*

***Item 12: Data Items: List and define all data points sought. Describe any assumptions made about any missing or unclear information.*| Mandatory | Modified from PRISMA-ScR**

Explanation: Data items refer to the specific units of information extracted from the included studies or records. When aggregated, these items provide the structured intelligence needed to generate insights and inform analysis.

*Rationale:* Clear reporting enables others to assess whether the chosen data items are appropriate, comprehensive, and aligned with the study objectives, while also revealing potential biases or gaps in the information considered. Transparency strengthens the credibility and reproducibility of the prioritisation process and ensures that stakeholders can trust that technologies were evaluated fairly and consistently.

*Example: “All technologies were further classified (see below) and all information is collated within the AI Dataset (Excel file accompanying this report). Classification of AI technologies:*

*• Clinical area and condition (if applicable/specified)*

*• Type of Scanning/ Medical Imaging (if applicable/specified) • Classification of technology by a) prevention and health promotion b) diagnosis and treatment c) intelligence operational automation • Country of Development*

*• Classification of development stage by Phase 1 (proof-of-concept stage); Phase 2 (prototype); Phase 3 (technology validated/demonstrated in relevant environment); Phase 4 (commercialised i.e. regulatory approved)*

*• Regulatory status/market authorisation (including list of approved markets) In addition to these fields, information related to clinical trials or published evidence was also captured. Furthermore, intelligence relating to funding/investment, company size, development awards or patents that was available during the review of sources was captured under ‘Additional Comments’ in the AI dataset (Excel file).”(Oyewole A et al., 2021)*

***Item 13: Effect measures:* *In line with the technology readiness level or technology type (medicine, diagnostic tests, medical devices or digital interventions), specify for each outcome the effect or performance measure(s) (e.g. risk ratio, mean difference, sensitivity or specificity, ROC, etc) used in the presentation of results.* | Mandatory | Modified from PRISMA-ScR**

**Explanation:** This item requires authors to clearly specify the effect or performance measures used to present outcomes for each technology identified in the horizon scan (e.g., sensitivity, specificity, risk ratio, ROC curves, or other relevant metrics depending on the technology type). Because horizon scans often include technologies at different readiness levels, the completeness and robustness of available data may vary—with early-stage innovations often lacking standardized or quantitative performance data. Acknowledging and reporting these variations is essential to provide appropriate context for interpretation, avoid overestimating the strength of evidence, and ensure transparency in how findings are presented.

**Rationale:** Including this information in the methods section of a horizon scan is crucial for ensuring clarity, transparency, and consistency in how results are interpreted. Defining effect or performance measures upfront explains how evidence will be evaluated for each technology type. Because technologies often differ in their stage of development, the quantity and quality of available data can vary widely. Recognizing and documenting these variations in the methods helps stakeholders assess the strength and limitations of the evidence, supports meaningful interpretation, and reinforces the methodological robustness and credibility of the horizon scan.

*Example: “Effect and performance measures were defined according to the type and maturity of each technology. For diagnostic tests, outcomes were reported using sensitivity, specificity, and ROC curves; for therapeutics, relative risk and mean difference were used where applicable. Given the variation in technology readiness levels across included innovations, the availability and completeness of performance data differed. Early-stage technologies often lacked standardized or quantitative measures. These differences were documented and considered when interpreting and presenting results.”*

***Item 14a: Synthesis methods:* *Describe the processes used to decide which signals were eligible for each analysis (item #7).* | Mandatory | Modified from PRISMA-ScR**

**Explanation:** This checklist item requires authors to clearly describe how signals identified during the horizon scan were assessed and selected for synthesis, ensuring a transparent and reproducible process. In the context of the HIPD/I framework—which incorporates *Horizon, Innovation, Population, and Data sources/Interest holders*—this involves specifying how inclusion and exclusion criteria were applied across these domains.

**Rationale:** Clearly outlining these criteria explains how the final set of signals was prioritized for analysis, supports consistency with the scan’s predefined scope, and enhances the transparency, validity, and strategic relevance of the findings.

*Example: “Signals identified during the initial search were assessed against predefined inclusion and exclusion criteria based on the HIPD/I framework. Specifically, innovations were included if they aligned with the defined horizon focus areas, represented a relevant innovation type (e.g., digital, diagnostic, therapeutic), addressed the target population of interest, and were supported by credible data sources or interest holder input. Signals that did not meet these criteria—such as those with insufficient evidence, limited relevance to the population, or falling outside the defined horizon—were excluded from synthesis* and listed in a separate appendix for transparency.**”**

***Item 14b: Synthesis methods:* *Describe any methods required to prepare the data for presentation or analyses, such as handling of missing data, or data conversions.* | Mandatory | Modified from PRISMA-ScR**

**Explanation:** This checklist item requires authors to explain how data were prepared for presentation or analysis, ensuring transparency and reproducibility. In a horizon scan, data often come from heterogeneous and incomplete sources, including early-stage evidence, stakeholder input, or grey literature. It is therefore important to describe any data processing methods used—such as cleaning and standardising information, handling missing or incomplete data, converting data into common formats, or categorising variables (e.g., by technology type or readiness level). Clear reporting of these steps helps readers understand how the data were structured to support synthesis, enables consistent interpretation, and strengthens the credibility of the findings.

**Rationale:** The rationale for this requirement in research that handles heterogeneous data is to ensure transparency, reproducibility, and validity of the analytical process. When data come from varied sources and formats—as is common in horizon scanning—preparation steps such as cleaning, standardising, and managing missing information can significantly influence the synthesis and interpretation of findings. By clearly documenting these methods, researchers allow others to understand, assess, and potentially replicate the process, reducing the risk of bias introduced through data handling decisions. This strengthens the robustness, credibility, and trustworthiness of the study’s conclusions.

*Example: “Data from multiple sources, including peer-reviewed literature, grey literature, and stakeholder submissions, were standardised before synthesis. Technology descriptions were harmonised using predefined categories (e.g., diagnostic, therapeutic, digital), and technology readiness levels were mapped to a common framework. Missing data on performance measures were recorded but not imputed; where appropriate, qualitative information was summarised narratively. All data were cleaned to remove duplicates and ensure consistency across sources.”*

***Item 15: Reporting bias assessment:*** ***If appropriate to the technology readiness level or technology type (medicine, diagnostic tests, medical devices or digital interventions) or scope of the scan, state if risk of bias of the studies used for identification of signals will be performed and which tools will be used.* | Optional | Modified from PRISMA-ScR**

**Explanation:** In the context of horizon scanning, this item refers to assessing the risk of bias in the sources or studies used to identify signals, where feasible. Because horizon scans often include early-stage technologies, grey literature, or expert input rather than fully developed clinical studies, it is not always possible or relevant to perform a formal bias assessment. When applicable—such as for more mature technologies with peer-reviewed evidence—authors should state whether a risk-of-bias assessment will be conducted and which tools (e.g., Cochrane Risk of Bias, ROBINS-I) will be used.

**Rationale:** Reporting this ensures transparency and credibility, helps stakeholders interpret the strength of the evidence underlying identified signals, and clarifies the limitations of the scan in relation to data quality.

*Example: “For this horizon scan, which focused on late-stage technologies, the risk of bias of the evidence supporting each signal was assessed where applicable. Peer-reviewed studies, including RCTs, cohort studies, and observational research, were evaluated using standard tools (Cochrane Risk of Bias 2 for RCTs and ROBINS-I for non-randomised studies). Evidence from press releases, regulatory announcements, and other grey literature sources was noted but not formally assessed for bias due to the nature of these sources. This approach allowed transparency regarding the strength and limitations of the evidence informing each identified signal.”*

***Item 16: Certainty assessment: If appropriate to the technology readiness level or technology type (medicine, diagnostic tests, medical devices or digital interventions), Describe any methods used to assess certainty (or confidence) in the body of evidence for an outcome using GRADE (Grading of Recommendations Assessment, Development, and Evaluation)* | Optional | Modified from PRISMA-ScR**

**Explanation:** In horizon scanning, this item involves assessing the certainty or confidence in outcomes reported across multiple technologies, particularly when the same outcome measure is reported for more than one innovation. It is important to note that a horizon scan is not intended to replace a systematic literature review, and the GRADE assessment in this context is typically based on a limited number of studies or sources. For early-stage technologies or data from expert opinion, press releases, or regulatory announcements, formal certainty assessment may not be feasible. Where evidence is available, tools like GRADE can provide a structured evaluation of reliability and consistency across technologies, helping stakeholders understand the strength of the evidence and its limitations. For these reasons, this item is optional and not mandatory in horizon scanning reports.

**Rationale:** Assessing the strength or certainty of evidence per outcome measure in horizon scanning allows stakeholders to understand how reliable the reported effects are across multiple technologies, which often vary in development stage and evidence quality. This enables prioritisation of technologies based on both potential impact and robustness of supporting evidence, highlights gaps where data are limited or inconsistent, and supports informed interpretation by acknowledging uncertainties. It also ensures transparency, helping decision-makers recognise the limitations of early-stage innovations and avoid overinterpreting preliminary findings, thereby strengthening the strategic and policy relevance of the scan.

*Example: “We applied GRADE to assess the certainty of evidence and to help prioritise technologies for implementation. GRADE is used to assess the strength of evidence for a specific outcome across studies, including nonclinical studies. Although most commonly used to assess evidence from a meta-analysis undertaken in the context of a systematic review, this framework can also be applied to individual studies or non-quantitative syntheses. We conducted GRADE assessments for three main outcomes for asthma: ‘asthma control’, ‘exacerbations’ and ‘diagnostic or prognostic accuracy’; and one main outcome for COPD: ‘prognostic value’.(Garcia Gonzalez-Moral et al., 2025)*

**Results**

Number of items in this section: 8

**Item 17a | *Weak signal selection: Describe the results of the search and selection process, from the number of records identified in the search to the number of technologies included in the horizon scan, ideally using a flow diagram* | Mandatory | Modified from PRISMA-ScR**

*Explanation: This item aligns closely with the PRISMA-ScR guidelines, as in both horizon scans and systematic reviews, the* number of records identified from each source during the search, how many of these records were screened, how many were excluded at each stage, and the final number of records included should be clearly presented. Ideally, this information should be presented in a flow diagram, outlining each step of the process, from searching to final selection. Horizon scans differ in that the records typically represent technologies rather than traditional study reports, and the focus of the report should reflect this.

*Rationale:* Providing this data ensures transparency, reproducibility, and demonstrates that a systematic approach was followed. It also presents information on the efficiency and size of the scan, helping to refine search strategies for future scans and inform on the resources needed. Additionally, it gives readers insight into the landscape of existing research and technologies, helping them understand trends and gaps in the field.

*Example:*

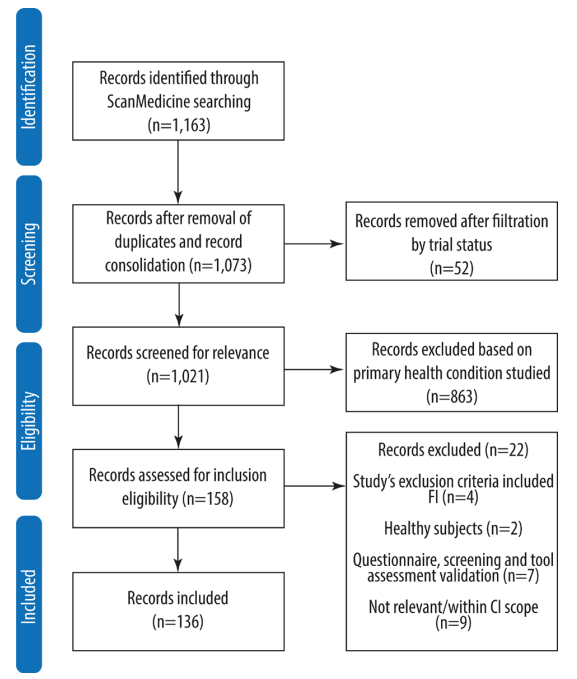

*A flow diagram from Thompson K, et al. 2022(Thomson et al., 2022)*

**Item 17b | *Weak signal selection: If appropriate include a list in appendix or separate tab of technologies that might appear to meet the inclusion criteria, but which were excluded, and explain why they were excluded. These may be referred to as ‘other interesting technologies’ that do not meet the criteria pre-established by stakeholders but present some innovative aspects of interest* | Optional | Modified from PRISMA-Scr**

*Explanation:* These could include technologies that meet some, but not all, of the inclusion criteria, or those for which insufficient data was available to determine eligibility for inclusion. They could also include technologies that, although not in remit, interest holders or other readers may be interested in.

*Rationale:* Horizon-scanning often explores areas where evidence is limited, and therefore including technologies that were considered, but ultimately excluded, helps to clearly define the boundaries and scope of the scan. It also ensures that promising or interesting technologies, which may evolve to become relevant in the future, are still documented. While including these technologies may risk inaccuracy, excluding them entirely can introduce bias. Providing a list of these technologies can help mitigate that bias.

*Example: “. The remaining deprioritised technologies are included in online supplemental appendix 3.”(Garcia Gonzalez-Moral et al., 2025)*

**Item 18 | *Single Source of information: If appropriate, cite each included single source of information and present its characteristics* | Mandatory | Modified from PRISMA-Scr**

*Explanation:* Horizon scans differ from systematic reviews as they aim to identify technologies rather than studies. The weak signals that inform on a single technology may come from multiple sources. Any single source of information is an individual source from where weak signals have been identified and data has been collected, and can include published or unpublished studies, a news item, funding call, patent documents or social media sources. Information on each single source should include source type and date of search. For grey literature, where and when it was accessed should be indicated. This information could be displayed in a table that could be added in appendix with links to external sources.

*Rationale:* This allows readers to assess the spread and the credibility of the sources and ensures the scan can be reproduced or built upon in future work.

*Example: “In November 2023 a comprehensive and systematic search for relevant information (within the Innovation Observatory’s ‘Medicines Innovation Database’ (MInD), ScanMedicine (a clinical trial database), global trial registries (e.g., ClinicalTrials.Gov, European Union Drug Regulating Authorities Clinical Trials Database (EudraCT), International Clinical Trials Registry Platform (ICTRP)), BiomedTracker (a site that provides pharmaceutical intelligence) was run.”(Wilkins & D., 2025)*

**Item 19 | *Technology characteristics: If appropriate for the scan scope, list technology characteristics* | Optional | New addition to checklist**

*Explanation:* Describe the technology details of each single source of information. Note that not all scans will aim to identify technology level characteristics as some scans may want to focus at clinical trial level. Reported technology characteristics should include stage of development, location, sponsor or originator, and technology details.

*Rationale: Clear and comparable reporting of technologies promotes full transparency of the results, supports clear interpretation by readers, and ensures the scan can be reproduced or built upon in future work.*

*Example: Headings in the results of a report by Jahan and Garcia Gonzalez-Moral (2025) include ‘Clinical Trial Landscape’, ‘Type of Technology’, ‘Development landscape’ (containing information about countries) and ‘Sponsor Information’.(Jahan A & Garcia Gonzalez-Moral S, 2024)*

**Item 20 | *Risk of bias in studies: If appropriate to the technology readiness level or technology type (medicine, diagnostic tests, medical devices or digital interventions), present assessments of risk of bias for each included study* | Optional | Modified from PRISMA-Scr**

*Explanation:* As horizon scans focus on early signals, not on evaluating the strength of effect size of evidence, assessing risk of bias (RoB) is optional and context dependent. The decision to perform a RoB assessment should weigh the time required against the value it adds to overall decision-making. RoB assessments may be more relevant for technologies in later stages of development, where clinical evidence begins to inform decisions. As different tools are suited to different study types, the most appropriate RoB approach should be selected if one is used. If no formal RoB assessment is performed, it is good practice, especially when comparing intervention outcomes, to comment on study design and key limitations.

*Rationale:* Although context dependent, assessing risk of bias, where appropriate, can strengthen the rigour and credibility of a horizon scan.

*Example: (An example of where limitations in study design, in this case reporting bias, are noted) “A significant challenge is the underreporting of generative AI model types, with 51% of technologies not disclosing this information. This poses challenges for regulators trying to assess these technologies in a timely manner given that different types of models can be associated with different levels of risks and types of mitigation requirements.”(Lanyi K et al., 2025)*

**Item 21 *| Results by intended purpose or outcome: If appropriate, present analyses by outcome, signal/s and/or technology/ies* | Optional | Modified from PRISMA-ScR**

*Explanation:* Present findings according to the intended purpose of the technology (e.g. diagnosis, treatment, monitoring, prevention), outcome (e.g. improved survival), signal type (e.g. clinical trial, funding, patent), or technology type (e.g. drug, device, diagnostic), depending on what best suits the scan’s objectives.

*Rationale:* This ensures the scan aligns with its objectives and can be tailored to decision-markers’ priorities, ultimately supporting clearer interpretation of the findings.

*Example: “As illustrated in Figure 4, the vast majority of technologies (68.8%) were developed to support diagnosis and treatment (including prognosis evaluation). 137 technologies (17.1%) were developed to aid in the prediction or prevention of a disease or health promotion. The remaining 113 technologies (14.1%) were developed to aid internal operations (intelligence operational automation).”(Oyewole A et al., 2021)*

**Item 22 *| Reporting missing data: Present missing data and how it has been dealt with in the analysis |* Mandatory | Modified from PRISMA-ScR**

*Explanation:* In horizon scans, data and data availability will change as technologies go through development, and so missing data needs to be transparently reported. It is also important to describe how missing data were handled in the analysis or summary. This may include approaches such as noting assumptions made, flagging technologies with particularly limited information, or highlighting areas where further monitoring is needed.

*Rationale:* Transparent handling of missing data not only helps readers interpret findings but also reinforces the scan’s accountability and replicability.

*Example: “Once the records for inclusion had been finalised, any missing data for variables which had been agreed in advance were supplemented using free text or categorical data available within other data sources.”(Wilkins & D., 2025)*

**Item 23 *| Certainty of evidence: If appropriate to the technology readiness level or technology type (medicine, diagnostic tests, medical devices or digital interventions), present assessments of certainty (or confidence) in the body of evidence for each outcome assessed. GRADE |* Optional | Modified from PRISMA-Scr**

*Explanation:* As horizon scans focus on early signals, assessing the certainty of evidence is optional and context dependent. It may be more relevant for technologies in later stages of development, where clinical evidence begins to inform decisions. When horizon scans compare intervention outcomes, it is good practice to comment on study design, key limitations, and the consistency or reliability of available data. However, unlike in systematic reviews, where formal grading systems like GRADE (Grading of Recommendations Assessment, Development, and Evaluation) are commonly used to evaluate certainty across well-defined outcomes from peer-reviewed studies, horizon scans often rely on diverse and incomplete data sources, such as preprints, news releases, patents, or early trial registrations. As such, formal certainty grading may not be feasible or meaningful in many cases, and so narrative or qualitative descriptions of confidence in the evidence should be used.(Brignardello-Petersen & Guyatt, 2025; Prasad, 2024)

*Rationale:* Although context dependent, assessing certainty of evidence, where appropriate, can strengthen the rigour and credibility of a horizon scan.

*Example: “Prioritisation: We applied GRADE to assess the certainty of evidence and to help prioritise technologies for implementation. GRADE is used to assess the strength of evidence for a specific outcome across studies, including nonclinical studies. Although most commonly used to assess evidence from a meta-analysis undertaken in the context of a systematic review, this framework can also be applied to individual studies or non-quantitative syntheses. We conducted GRADE assessments for three main outcomes for asthma: ‘asthma control’, ‘exacerbations’ and ‘diagnostic or prognostic accuracy’; and one main outcome for COPD: ‘prognostic value’.”(Garcia Gonzalez-Moral et al., 2025)*

**Discussion**

Number of items in this section: 4

**Item 24a | *Discussion: Provide a general interpretation of the results in the context of other Political, Economic, Social, Technological, Legal, and Environmental (PESTLE) factors* | Mandatory | Modified from PRISMA-Scr**

*Explanation:* Identify how Political, Economic, Social, Technological, Legal, and Environmental (PESTLE) factors shape the relevance, feasibility, or uptake of the innovations identified. Also, can be used to provide insight into why particular technologies are being developed or prioritised.

*Rationale:* This is introduced here to align with the PESTLE framework, a common tool in horizon-scanning that considers Political, Societal, Technological, Legal, and Environmental factors. This helps identify gaps in research, anticipate potential enablers or barriers to technology development and adoption, and provides context to the scan.

*Example of economic factors affecting innovation: “Our results showed little evidence of new pharmaceutical technologies in development … This is perhaps not surprising considering the vast costs of developing new technologies … Given the vast costs associated with research and development, pharmaceutical companies are perhaps unwilling to invest in faecal incontinence innovations.”(Thomson et al., 2022)*

**Item 24b | *Discussion: Discuss any limitations of the data included in the horizon scan |* Mandatory | Modified from PRISMA-Scr**

*Explanation:* Examples of data limitations in a horizon scan include incomplete, preliminary, or fragmented data, publication bias, and potential bias caused unvalidated or non-peer-reviewed sources

*Rationale:* Horizon scans rely on a mix of early-stage, non-peer-reviewed sources such as trial registries, press releases, and patents. The limitations associated with these sources can affect confidence in the signals identified and their potential future impact, and so commenting on them supports informed interpretation.

*Example: “However, other relevant clinical trials may not have been identified in this process if their registration records were not available through ScanMedicine. Some of the identified trial records were not recently updated, making it difficult to assess their current state of development, or whether they had been prematurely discontinued.”(Thomson et al., 2022)*

**Item 24c | *Discussion: Discuss any limitations of the horizon-scanning processes used (inc. language barriers - only English)* | Mandatory | Modified from PRISMA-Scr**

*Explanation:* Examples of limitations of the horizon-scanning processes include time or resource constraints that limit searches, and selection bias, such as focusing only on English-language sources, prioritising certain databases over others, or relying on sources that are more accessible.

*Rationale:* Commenting on these limitations supports transparency, defines the limit of the scan’s findings, and helps identify areas where further evidence is needed.

*Example: “The process we used to horizon scan for this project focuses on products that have gone through clinical trials and may have missed relevant products that do not require a review of efficacy and safety as part of the regulatory processes...”(Thomson et al., 2022)*

**Item 24d | *Discussion: Discuss implications of the results for practice, policy, and future research* | Mandatory | Unmodified from PRISMA-Scr**

*Explanation:* Reflect on how the findings might inform decision-making, implementation planning, or investment in healthcare practice and policy. Consider whether any of the identified technologies suggest emerging trends that require regulatory, clinical, or resource readiness. Additionally, identify areas where further research may be needed.

*Rationale:* This helps inform next steps and ensures that results are translated into meaningful actions, increasing the impact of the horizon scan.

*Example: “… there is a clear justification to move the focus to effective combination therapies. Focusing on safe and effective combinations in future randomised controlled trials may enable better overall therapeutic efficacy against COVID-19. Large, randomised combination therapy clinical trials are warranted using some interventions with significant, albeit small, benefits in monotherapy trials rather than continued duplication of ongoing trials … Combination therapies can also be evaluated in platform trials … This will enable early prioritisation of effective treatment arms while saving time and resources.”(Akinbolade et al., 2022)*

**Other Information**

Number of items in this section 6.

**Item 25a** **| *Registration and protocol: Provide registration information for the review, including register name and registration number, or state that the review was not registered.* | Mandatory | Unmodified from PRISMA-ScR**

**Explanation:** This item requires authors to clearly state whether the review was registered in a publicly accessible database (e.g., Open Science Framework, PROSPERO if appropriate), and if so, to provide the register name and registration number. If the review was not registered, this must also be explicitly stated.

**Rationale:** Providing registration information in a horizon scan enhances transparency and accountability by documenting the planned scope, methods, and timelines. It helps minimize bias, supports reproducibility, and enables others to identify ongoing or related scans, reducing duplication of effort. Clearly stating whether the scan is registered - along with the register name and number if applicable - aligns with good reporting practices and strengthens the credibility and integrity of the work.

Example: *“This horizon scan was registered with [Register Name] (Registration Number: [XXXXXX]) prior to initiation. The protocol, including scope, objectives, and methods, is publicly accessible at [link]. If the scan was not registered, this could be stated as: “This horizon scan was not prospectively registered on a publicly accessible register.”*

**Item 25b | *Registration and protocol: Indicate where the review protocol can be accessed, or state that a protocol was not prepared.* | Mandatory | Unmodified from PRISMA-ScR**

**Explanation:** In addition to item 25a, this item aims to provide additional protocol information.

**Rationale:** For a horizon scan, this item ensures transparency by making the planned scope and methods publicly available, helps reduce bias, allows others to identify overlapping scans to avoid duplication, and supports reproducibility and credibility through clear documentation of the original plan.

*Example: “The protocol for this horizon scan was prospectively registered on the Open Science Framework (OSF; Registration Number: [XXXXXX]) and is available at [link]. This registration outlines the objectives, scope, and methodological approach. If not registered: ‘This horizon scan was not registered on a publicly accessible platform.”*

**Item 25c | *Registration and protocol: Describe and explain any amendments to information provided at registration or in the protocol.* | Mandatory | Unmodified from PRISMA-ScR**

**Explanation:** This item means authors should report and explain any changes made to the original plan or protocol after registration or initial documentation. This ensures transparency, allows readers to understand deviations from the planned scope or methods, and supports the credibility and reproducibility of the scan.

**Rationale:** This is to ensure transparency and maintain trust in the research process. By documenting and explaining any amendments to the registered protocol or initial plan, readers can see how and why the study deviated from its original design. This helps assess the risk of bias, understand the context of the findings, and supports reproducibility and accountability. Without such reporting, changes could obscure selective reporting or methodological shifts that affect the interpretation of results.

Example: *“Since the initial registration of this horizon scan on the Open Science Framework (OSF; Registration Number: [XXXXXX]), minor amendments were made to the search strategy and inclusion criteria to better capture emerging technologies. These changes are documented in the updated protocol available at [link], along with a rationale for each amendment.”*

**Item 26 | *Support: Describe sources of financial or non-financial support for the review, and the role of the funders or sponsors in the review.* | Mandatory | Unmodified from PRISMA-ScR**

**Explanation:** Reporting support in a horizon scan is important for transparency and credibility. It allows readers to understand who funded or otherwise supported the work and whether these entities had any influence over the design, scope, methodology, or interpretation of findings. Clearly stating the role of funders or sponsors helps identify potential conflicts of interest, reassures readers that the scan was conducted independently, and aligns with good scientific and ethical reporting practices.

**Rationale:** The rationale for reporting funding or support in research, including horizon scanning, is to enhance transparency, accountability, and trustworthiness. Funding sources or sponsors can introduce potential conflicts of interest or biases, consciously or unconsciously influencing the scope, methodology, or interpretation of findings. In horizon scanning, where the identification of emerging technologies or trends can inform policy or strategic decisions, disclosing support ensures that stakeholders can assess the independence of the scan, the credibility of the findings, and the reliability of any recommendations derived from the work.

*Example:* *“This horizon scan was supported by [Funding Organization] through a research grant (Grant Number: [XXXX]). The funders had no role in defining the scope, methodology, data collection, analysis, or interpretation of findings, ensuring that the scan was conducted independently.”*

**Item 27 | *Competing interests: Declare any competing interests of review authors.* | Mandatory | Unmodified from PRISMA-ScR**

**Explanation:** In a horizon scanning report, declaring competing interests requires authors to disclose any financial, professional, or personal relationships that could influence, or be perceived to influence, the findings or interpretation of the scan.

**Rationale:** This is important to maintain transparency and credibility, allowing readers to assess potential bias and trust the independence of the assessment, particularly when the scan may inform policy, strategy, or investment decisions.

*Example: “The authors declare that they have no competing interests relevant to this horizon scan.” Or, if relevant interests exist: “Author A has received consultancy fees from [Company/Organization], which is involved in [relevant field]. All other authors declare no competing interests. These relationships did not influence the design, conduct, or reporting of this horizon scan.”*

**Item 28 | *Availability of data, code and other materials: Subject to agreement with stakeholder, report which of the following are publicly available and where they can be found: template data collection forms; data extracted from included studies; data used for all analyses; analytic code; any other materials used in the review.* | Mandatory | Modified from PRISMA-ScR**

**Explanation:** PRISMA-ScR item requires authors to clearly report what materials from the review are publicly accessible and where they can be obtained. This includes data collection forms, extracted data, analytic datasets, code, or other materials used in the review. The purpose is to enhance transparency, reproducibility, and reusability of the research, allowing others to verify findings, replicate analyses, or build upon the work. In cases where materials cannot be shared due to stakeholder agreements or confidentiality, this should be explicitly stated**.**

**Rationale:** In horizon scanning, like in scoping reviews, sharing data and materials promotes transparency, reproducibility, and credibility, allowing others to verify findings or adapt methods, while explicitly noting restrictions maintains accountability.

*Example: “All data collection templates, extracted data, and analysis code used in this horizon scan are publicly available at [repository link]. Some materials containing sensitive stakeholder information are not shared due to confidentiality agreements.”*

**References:**

Akinbolade, S., Coughlan, D., Fairbairn, R., McConkey, G., Powell, H., Ogunbayo, D., & Craig, D. (2022). Combination therapies for COVID-19: An overview of the clinical trials landscape. *Br J Clin Pharmacol*, *88*(4), 1590-1597. <https://doi.org/10.1111/bcp.15089>

Brignardello-Petersen, R., & Guyatt, G. H. (2025). Assessing the certainty of the evidence in systematic reviews: importance, process, and use. *Am J Epidemiol*, *194*(6), 1681-1686. <https://doi.org/10.1093/aje/kwae332>

Garcia Gonzalez-Moral, S., Addis, P., Sharma, O., Oliver, A., Johnson, E. E., Al-Assaf, A., Sadiq, A., & Meader, N. (2025). Innovative technologies for asthma and COPD management in the community: scanning the horizon using rapid systematic review methods. *BMJ Innovations*, bmjinnov-2024-001330. <https://doi.org/10.1136/bmjinnov-2024-001330>

Hines, P., Hiu Yu, L., Guy, R. H., Brand, A., & Papaluca-Amati, M. (2019). Scanning the horizon: a systematic literature review of methodologies. *BMJ Open*, *9*(5), e026764. <https://doi.org/10.1136/bmjopen-2018-026764>

Jahan A, & Garcia Gonzalez-Moral S. (2024). *Horizon Scanning Report: Summary of horizon scans and landscape analyses of innovations in antimicrobial resistance technologies*. <https://io.nihr.ac.uk/wp-content/uploads/2024/11/AMR-Report_Sept-2024_.pdf>

Oparah C, Eastaugh C, Lanyi K, Hussain A, Woltmann J, Pearson F, Still M, Briush E, & N, K. (2023). *Interim Horizon Scanning Report: Identification of paediatric neurological trauma technologies*. <https://io.nihr.ac.uk/wp-content/uploads/2023/11/NIHRIO-MDx-Scan-Report-Paediatric-Trauma_Oct-2023.pdf>

Oyewole A, Marshall C, Robertson E, Alipat M, Dangova K, Sarpal H, Barrass L, & Garcia Gonzalez-Moral, S. (2021). *Final Report: Identification of AI technologies in Healthcare*. <https://www.io.nihr.ac.uk/wp-content/uploads/2023/04/050221-NIHRIO-AI-Scan-Report-v1.pdf>

Prasad, M. (2024). Introduction to the GRADE tool for rating certainty in evidence and recommendations. *Clinical Epidemiology and Global Health*, *25*, 101484. <https://doi.org/https://doi.org/10.1016/j.cegh.2023.101484>

Thomson, K. H., Dangova, K., Bliss, D. Z., Wallace, S., O'Connor, N., Richter, H. E., & Pearson, F. (2022). Future developments and new technologies in the field of faecal incontinence: scanning the horizon using late-stage clinical trial registrations. *BMJ Innovations*, *8*(4), 278-284. <https://doi.org/10.1136/bmjinnov-2021-000860>

Tricco, A., Lillie, E, Zarin, W, O'Brien, KK, Colquhoun, H, Levac, D, Moher, D, Peters, MD, Horsley, T, Weeks, L, Hempel, S et al. (2018). PRISMA Extension for Scoping Reviews (PRISMA-ScR): Checklist and Explanation. *Annals of Internal Medicine*, *169*(7), 467-473. <https://doi.org/10.7326/m18-0850> %m 30178033

Wilkins, G., Potter, R., Ewedairo, O., Nesworthy, J., Pennock, E., Williams, O., O’Keefe, H., Craig,, & D., P., F., & Mkwashi, A. (2025). *A pipeline analysis of healthcare technologies in development for addiction*. <https://doi.org/10.6084/m9.figshare.28771112>
