## Supplementary material for "How should horizon scanning studies be reported? Developing a checklist of standard items": Horizon Scanning checklist

| Section and Topic | Item | Checklist item | Page Number |
| --- | --- | --- | --- |
| TITLE | | |  |
| Title | 1 | Identify the report as a horizon scan or other alternative terms used to refer to horizon scanning outputs for example landscape or pipeline analyses/report or broader umbrella terms such as foresight analysis |  |
| ABSTRACT | | |  |
| Abstract | 2 | See adaptation of the PRISMA 2020 for Abstracts checklist for horizon scanning reports. |  |
| INTRODUCTION | | |  |
| Rationale | 3 | Describe the rationale for the horizon scan in the context of existing knowledge, unmet needs, policy or emerging innovation. |  |
| Interest holder | 4 | Disclosure of interest holder name and brief function or role in decision-making |  |
| Objectives | 5 | Provide an explicit statement of the objective(s) or question(s) the horizon scan addresses. |  |
| METHODS | | |  |
| Scope | 6 | Consider using the HIP-D/I framework: Horizon, Innovation, Population – Data source/ Interest Holder concepts |  |
| Eligibility criteria | 7 | Specify the inclusion and exclusion criteria for the horizon scan |  |
| Information sources | 8 | Specify all databases, registers, websites, organisations, reference lists, PPIE and expert solicitation and consultation as well as any other sources searched or consulted to identify signals. Specify the date when each source was last searched or consulted. |  |
| Search strategy | 9 | Present the full search strategies for all databases, registers and websites, including any filters and limits used. Report if any AI tool has been used for creating or running the searches and provide name and version number of the tool. |  |
| Selection process | 10 | Specify the methods used to decide whether a signal met the inclusion criteria of the horizon scan, including how many reviewers screened each record and each report retrieved, whether they worked independently, and if applicable, details of automation tools used in the process. |  |
| Data collection process | 11 | Specify the methods used to collect data from reports, including how many reviewers collected data from each report, whether they worked independently, any processes for obtaining or confirming data from third parties, and if applicable, details of automation tools used in the process. |  |
| Data items | 12 | List and define all data points sought. Describe any assumptions made about any missing or unclear information. |  |
| Effect measures | 13 | In line with the technology readiness level or technology type (medicine, diagnostic tests, medical devices or digital interventions), specify for each outcome the effect or performance measure(s) (e.g. risk ratio, mean difference, sensitivity or specificity, ROC, etc) used in the presentation of results. |  |
| Synthesis methods | 14a | Describe the processes used to decide which signals were eligible for each analysis (item #7). |  |
|  | 14b | Describe any methods required to prepare the data for presentation or analyses, such as handling of missing data, or data conversions. |  |
| Reporting bias assessment | 15 | If appropriate to the technology readiness level or technology type (medicine, diagnostic tests, medical devices or digital interventions) or scope of the scan, state if risk of bias of the studies used for identification of signals will be performed and which tools will be used. |  |
| Certainty assessment | 16 | If appropriate to the technology readiness level or technology type (medicine, diagnostic tests, medical devices or digital interventions), Describe any methods used to assess certainty (or confidence) in the body of evidence for an outcome using GRADE (Grading of Recommendations Assessment, Development, and Evaluation) |  |
| RESULTS | | |  |
| Weak signal selection | 17a | Describe the results of the search and selection process, from the number of records identified in the search to the number of technologies included in the horizon scan, ideally using a flow diagram. |  |
|  | 17b | If appropriate include a list in appendix or separate tab of technologies that might appear to meet the inclusion criteria, but which were excluded, and explain why they were excluded. These may be referred to as ‘other interesting technologies’ that do not meet the criteria pre-established by interest holders but present some innovative aspects of interest. |  |
| Single source of information characteristics | 18 | Cite each included single source of information and present its characteristics. |  |
| Technology characteristics | 19 | If appropriate for the scan scope, list technology characteristics. |  |
| Risk of bias in studies | 20 | If appropriate to the technology readiness level or technology type (medicine, diagnostic tests, medical devices or digital interventions), present assessments of risk of bias for each included study. |  |
| Results by intended purpose or outcome | 21 | If appropriate, present analyses by outcome, signal/s and/or technology/ies. |  |
| Reporting missing data | 22 | Present missing data and how it has been dealt with in the analysis. |  |
| Certainty of evidence | 23 | If appropriate to the technology readiness level or technology type (medicine, diagnostic tests, medical devices or digital interventions), present assessments of certainty (or confidence) in the body of evidence for each outcome assessed using GRADE (Grading of Recommendations Assessment, Development, and Evaluation). |  |
| DISCUSSION | | |  |
| Discussion | 24a | Provide a general interpretation of the results in the context of other Political, Economic, Social, Technological, Legal, and Environmental (PESTLE) factors |  |
|  | 24b | Discuss any limitations of the data included in the horizon scan. |  |
|  | 24c | Discuss any limitations of the horizon scanning processes used (inc. language barriers - only English). |  |
|  | 24d | Discuss implications of the results for practice, policy, and future research. |  |
| OTHER INFORMATION | | |  |
| Registration and protocol | 25a | Provide registration information for the review, including register name and registration number, or state that the review was not registered. |  |
|  | 25b | Indicate where the review protocol can be accessed, or state that a protocol was not prepared. |  |
|  | 25c | Describe and explain any amendments to information provided at registration or in the protocol. |  |
| Support | 26 | Describe sources of financial or non-financial support for the review, and the role of the funders or sponsors in the review. |  |
| Competing interests | 27 | Declare any competing interests of review authors. |  |
| Availability of data, code and other materials | 28 | Subject to agreement with stakeholder, report which of the following are publicly available and where they can be found: template data collection forms; data extracted from included studies; data used for all analyses; analytic code; any other materials used in the review. |  |

Shaded rows (grey) – optional items; Red writing – newly introduced or modified items
